# Developing and validating a multivariable prediction model for predicting costs of colon surgery

**DOI:** 10.1101/2022.02.02.22270329

**Authors:** Anas Taha, Stephanie Taha-Mehlitz, Vincent Ochs, Bassey Enodien, Michael Drew Honaker, Daniel M. Frey, Philippe C. Cattin

## Abstract

Hospitals are burdened with predicting, calculating and managing various cost-affecting parameters regarding patients and their treatments. Accuracy in cost prediction is further affected if a patient suffers from other health issues which hinder the traditional prognosis. This can lead to an unavoidable deficit in the final revenue of medical centers. This study aims to determine whether machine learning (ML) algorithms can predict cost factors based on patients undergoing colon surgery. For the forecasting, multiple predictors will be taken into the model to provide a tool that can be helpful for hospitals to manage their costs which ultimately will lead to operating more cost-efficiently.. This proof of principle will lay the groundwork for an efficient ML-based prediction tool based on multicenter data from a range of international centers in the subsequent phases of the study. With a % MAPE result of 18 – 25.6, our model’s prediction showed decent results to forecast the costs regarding various diagnosed factors and surgical approaches. There is an urgent need for further studies on predicting cost factors, especially for cases with anastomotic leakage, to minimize unnecessary costs for hospitals.

## 1. Introduction

### 1.1 Background

Colorectal cancer (CRC)is one of the most prevalent cancers in the world today based on diagnoses, with about 1.8 million cases being diagnosed and about 0.7 million related deaths occurring annually. In addition, CRC accounts for 10 % of all newly diagnosed cancers, a considerable social and economic burden for many nations worldwide (1). One of the treatment modalities for colorectal cancer is surgery. Surgery is aimed at obtaining an adequate oncologic resection while re-establishing intestinal continuity. Over time, there have been improvements in the way the disease is treated. But existing patien comorbidities can limit the surgical procedures. The time required to prepare patients for surgery and address their comorbidities contribute to increased surgical costs. However, despite many improvements, significant other complications still occur during, and especially after, a surgical procedure. To avoid this, the patient is placed in necessary post-operative care, generally for 5 and 7 days after a surgical operation. Other post-operative risk factors will further add to the surgical cost, but their prediction is very vague due to the absence of sufficient datasets. These involve performing a colorectal anastomosis, anastomotic leak, (2), delirium or prolonged ileus (3), other emergency surgeries, longer intra-operative time, and peritoneal contamination

The comorbidities and longer stays result in a cost burden for patients and hospitals. This is why prediction models are now being updated to determine the costs for anastomotic insufficiency. Prediction models are normally used to estimate the probability of achieving a particular outcome (4). A large number of prediction models have been developed, but only a small number are used because not all models accurately predict the desired outcome (5). This study focuses on developing and validating a multivariable prediction model to predict costs for patients undergoing colon surgery while considering their stay in the hospital. This will help determine the cost burden due to variable hospital length of stay (LOS) as well as days spent in intensive care units (ICU). The medical context is prognostic in that it is focused on predicting the cost of overall expenditure involved in colon surgery for the clinical center and the patient.

### 1.2 Rationale

The rationale for developing and validating the multivariable model is that it will help accurately predict the costs associated with colon surgery. Accurate prediction will help patients and practices employeed by the hospital make more informed decisions, as well as aid in policies enacted by the government. The results that come with the use of the model will also aid in surgical planning. In short, developing and validating the multivariable model will provide insight into costs involved in colon surgery. In turn, it will allow revisions in care and help develop strategies for improved management. Similar studies for prediction purposes have been conducted in the field of medicine. For example, Musunuri et al. have used machine learning in the form of artificial intelligence to predict 90-day liver disease mortality. Focused on acute-on-chronic liver failure, they achieved a model with an accuracy of 94.12% and an area under the curve of 0.915 (6). Hameed et al. wrote about the impact of artificial intelligence on urological diseases. In their literature review, they have pointed to multiple publications using various models like support vector machine, nearest neighbour, random forrest, convolutional neural network or artificial neural networks to predict and classify diseases like prostate cancer, urothelial cancer, renal cancer or urolithiasis. What differs from those publications and their work from ours is, that they use a classification model instead of a regression model. The most important benefit of using a regression model compared to a classification model is that it helps predict continuous values, whereas classification models try to predict discrete class labels. To predict the costs associated with colon surgery in an accurate way, a machine learning regression model is used. Using this approach. We aim to contribute to an existing gap in this field (7).

### 1.3 Objectives

- To develop prediction models for the final costs in patients based on multiple predictors.
- To test the models in terms of their ability to accurately predict the final costs associated with colon surgery in patients.

## 2. Methods

### 2.1 Overview and Data Collection

Data was extracted from a registry of patients who underwent colonic anastomosis for various reasons such as tumors, diverticulitis, mesenterial ischemia, iatrogenic or traumatic perforation, or inflammatory bowel disease (aggregated as “non-tumor”) at the Hospital of Wetzikon from 1^st^ January 2013 to the 31^st^ December of 2019. No patients were excluded from the initial data collection. Furthermore, this study was completed based on the transparent reporting of a multivariable prediction model for individual prognosis or diagnosis (TRIPOD) statement checklist for prediction model development (8).

Utilizing this data, we developed a machine learning model to predict the costs of colon surgery.

### 2.2 Ethical Considerations

The registry data was approved by an institutional review board, where the patients’ informed consent was waived. The study was registered at [Req 2021-01107].

### 2.3 Predictors and Outcome Measures

Recorded variables include Insurance (general/semi-private/private), age, surgical procedure (Hartmann/left sided hemicolectomy and extended left sided hemicolectomy/ right sided hemicolectomy and extended right sided hemicolectomy/sigmoid resection), surgical approach (open/laparoscopic), diagnosis (tumor/non-tumor), final costs (the sum of all cost factors), length of stay (in days), Intensive care stay (in days), operation time (in minutes), anaesthesia time (in minutes), ASA-Score (I,II,III,IV), gender (male/female), CCI (Charlson Comorbidity Index), anastomotic insufficiency and emergent/non-emergent. The data on the final cost, which is the sum of all the costs incurred during the stay in hospital for surgery, were collected in CHF (Swiss Francs). Other cost factors not incorporated since they add up to the final costs include administrative costs, costs of hospitality, nurse costs, costs of infrastructure, doctor costs, medical costs, operational costs, anesthesia costs and care costs.

### 2.4 Model Development

Data was randomly split into two sets, 80% of the data was put into a training set to build the models and 20% was utilized for a test set to validate the models and assess their performance internally. The two sets had approximately the same class distribution (Gaussian). The following 14 predictors were chosen to predict the final costs based on regression and clinical insights: age, gender, insurance, diagnosis, operation, surgical approach, hospitalization, intensive care, surgical procedure and anaesthesia time, CCI, ASA-Score, anastomotic insufficiency and emergency surgery (9).

By having variables included, such as the CCI and the ASA-Score, we are able to cover a large number of diseases that are included in the comorbidity index.

A variety of machine learning models were developed, including generalized boosted regression, random forest, and decision trees. An interaction depth of 3 and a total number of 500 trees were chosen, as was the type of the random forest and the regression model. The classification/predictive performance was measured using the mean absolute percentage error (MAPE), where a result of < 10% is classified as highly accurate, <20% denotes as a good forecast 20%-50% as a reasonable forecast and everything >50% as an inaccurate forecast (10). The MAPE factor, also known as mean absolute percentage deviation (MAPD), is used for accuracy of a forecasting prediction. Continuous data are reported as mean ± standard deviation (SD) or median (interquartile range (IQR)) and categorical data as numbers (percentages). Hyperparameters were tuned, and the final model was selected based on the MAPE. The final model chosen was the random forest model based on its superior performance.

The analysis was carried out using R version 4.0.4. The random forest library was used for the random forest models, the metrics library used for the calculation of the performance measurements, the gbm library for the generalized boosted regression models and the rpart library was used for the other models.

### 2.5 Deployment

The best performing model will be deployed as a web-based, user-friendly application using RShiny to predict the final costs, that considers the different cost factors.

(Accessible via: https://colonsurgerycost.shinyapps.io/Final_Cost/).

## 3. Results

### 3.1 Cohort

A total of 347 patients were included in our study. This number consists of all patients from the center who suffered from the diagnosed factors of section 3 and had to undergo the type of operations mentioned. The mean age was 67 ± 14 years (range 28-94). 162 (47%) patients were male and 185 (53%) were female. Tables 1 and 2 provide all baseline variables and their descriptive statistics. Continuous variables were recorded as mean ± SD (range) in Table 1.

**Table 1.**
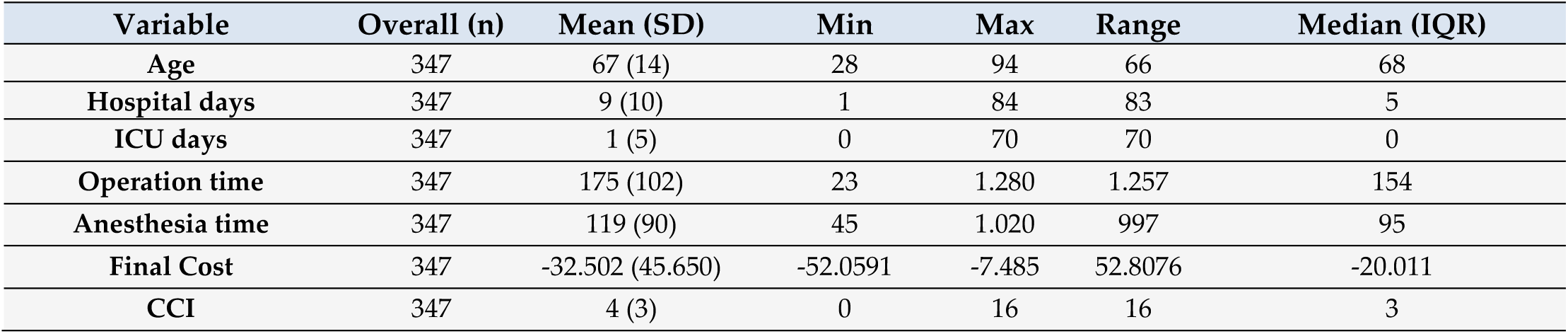
Variable characteristics for continuous values.

| Variable | Overall (n) | Mean (SD) | Min | Max | Range | Median (IQR) |
| --- | --- | --- | --- | --- | --- | --- |
| Age | 347 | 67 (14) | 28 | 94 | 66 | 68 |
| Hospital days | 347 | 9 (10) | 1 | 84 | 83 | 5 |
| ICU days | 347 | 1 (5) | 0 | 70 | 70 | 0 |
| Operation time | 347 | 175 (102) | 23 | 1.280 | 1.257 | 154 |
| Anesthesia time | 347 | 119 (90) | 45 | 1.020 | 997 | 95 |
| Final Cost | 347 | -32.502 (45.650) | -52.0591 | -7.485 | 52.8076 | -20.011 |
| CCI | 347 | 4 (3) | 0 | 16 | 16 | 3 |

**Table 2.** Variable characteristics for categorical values.

| Variable | n (%) |
| --- | --- |
| Gender |  |
| Male | 162 (47%) |
| Female | 185 (53%) |
| **Insurance** |  |
| General | 283 (82%) |
| Semi-Private | 49 (14%) |
| Private | 15 (4%) |
| **Diagnosis** |  |
| Tumor | 162 (47%) |
| Non-Tumor | 185 (53%) |
| **Emergency Surgery** |  |
| No | 331 (95%) |
| Yes | 16 (5%) |
| **Operation** |  |
| Hartmann`s procedure | 19 (5%) |
| Hemicolectomy left | 16 (4%) |
| Extended Hemicolectomy left | 6 (2%) |
| Hemicolectomy right | 82 (24%) |
| Extended Hemicolectomy right | 6 (2%) |
| Sigmoid resection | 218 (63%) |
| **Surgery approach** |  |
| Open | 153 (44%) |
| Laparoscopic | 194 (56%) |
| **Anastomotic insufficiency** |  |
| No | 331 (95%) |
| Yes | 16 (5%) |
| **ASA Score** |  |
| I | 12 (4%) |
| II | 184 (53%) |
| III | 137 (39%) |
| IV | 14 (4%) |

Categorical variables were recorded as numbers (percentage) on Table 2. No missing values were detected. Table 3 provides the variables’ characteristics and descriptive statistics that are not mentioned in Table 1 or Table 2 and are based on their impact on the final costs.

**Table 3.** Descriptive statistics based on final costs.

| Variable | Overall (n) | Mean (SD) | Min | Max | Median (Q1, Q3) | P-Value |
| --- | --- | --- | --- | --- | --- | --- |
| **Insurance** | 347 | -32.502 (45.650) | -520.591 | -7.485 | -20.011 (-28.828, -15.332) | 0.643 |
| General | 283 | -31.773 (47.490) | -520.591 | -7.485 | -18.433 (-27.464, -14.823) |  |
| Semi-Private | 49 | -33.495 (27.892) | -192.811 | -10.919 | -22.645 (-40.043, -19.795) |  |
| Private | 15 | -42.996 (57.232) | -241.331 | -13.915 | -23.979 (-39.017, -18.426) |  |
| **Diagnosis** | 347 | -32.502 (45.650) | -520.591 | -7.485 | -20.011 (-28.828, -15.332) | 0.842 |
| Tumor | 162 | -33.025 (40.120) | -298.957 | -7.485 | -21.129 (-28.294, -15.790) |  |
| Non-Tumor | 185 | -32.043 (50.098) | -52.059 | -9.929 | -19.155 (-29.688, -14.859) |  |
| **Operation** | 347 | -32.502 (45.650) | -52.0591 | -7.485 | -20.011 (-28.828, -15.332) | <0.001 |
| Hartmann | 19 | -25.479 (20.230) | -75.676 | -7.485 | -18.433 (-24.551, -14.053) |  |
| Hemicolectomy left | 16 | -65.777 (91.964) | -38.419 | -15.045 | -32.297 (-70.713, -18.876) |  |
| Extended<br>Hemicolectomy left | 6 | -11.0698 (20122) | -520.591 | -13.915 | -28.751 (-46.173, -22.338) |  |
| Hemicolectomy right | 82 | -35135 (39.474) | -241.331 | -10.665 | -22.469 (-35.663, -16.468) |  |
| Extended<br>Hemicolectomy right | 6 | -65.768 (114.464) | -298.957 | -13.086 | -17.726 (-29.401, -14.799) |  |
| Sigmoid resection | 218 | -26.613 (23.764) | -192.811 | -9.379 | -18.684 (-25.538, -15.180) |  |
| **Surgery approach** | 347 | -32.502 (45.650) | -520.591 | -7.485 | -20.011 (-28.828, -15.332) | <0.001 |
| Open | 153 | -46.531 (64.486) | -520.591 | -7.485 | -25.989 (-45.758, -18.708) |  |
| Laparoscopic | 194 | -21.437 (13.486) | -91.098 | -9.379 | -17.275 (-21.765, -14.685) |  |
| **Anastomotic**<br>**insufficiency** | 347 | -32.502 (45.650) | -520.591 | -7.485 | -20.011 (-28.828, -15.332) | <0.001 |
| No | 331 | -26.051 (20.763) | -192.811 | -7.485 | -19.472 (-27.204, -15.121) |  |
| Yes | 16 | -165.941 (136.653) | -520.591 | -27.444 | 114.158 (225.666, -78.015) |  |
| **ASA Score** | 347 | -32.502 (45.650) | -520.591 | -7.485 | -20.011 (-28.828, -15.332) | <0.001 |
| I | 12 | -20.626 (5.177) | -30.208 | -14.035 | -20.982 (-23.627, -16.212) |  |
| II | 184 | -23.129 (21.680) | -241.331 | -7.485 | -17.938 (-22.591, -14.515) |  |
| III | 137 | -38.328 (44.857) | -384.159 | -10.665 | -22.645 (-43.274, -16.997) |  |
| IV | 14 | -108.844 (140.598) | -520.591 | -20.280 | -53.515 (-79.035, -33.144) |  |

### 3.2 Model Performance

During internal validation, the performance of all three models was tested and stated with their mean value and their 95% confidence intervals (Table 3). The random forest classifier provided the highest MAPE for predicting the final costs (21.4). Thus, it was the model with the best internal validation performance and was subsequently used for predicting costs (11). In comparison, the decision tree and general boosted regression model displayed results for MAPE of only 25.5 and 29.7, respectively. Therefore, the average of the MAPE for the final costs is around 21.4 which means that on average, the forecast of this prediction model regarding the final costs are off by 21.4%. Since a MAPE value of <20% is considered as being “good”, our result is showing decent results. The percentage of the random forest classifier’s variance, which was explained in the models, varied from 73.81% to 81.05%. Specific feature importance according to the random forest classifier is displayed as Gini Index in Figure 1, while Figure 2 shows the prediction of the Random Forest Classifier compared to the actual observed values from the test data set for the final cost factor.

**Figure 1.**
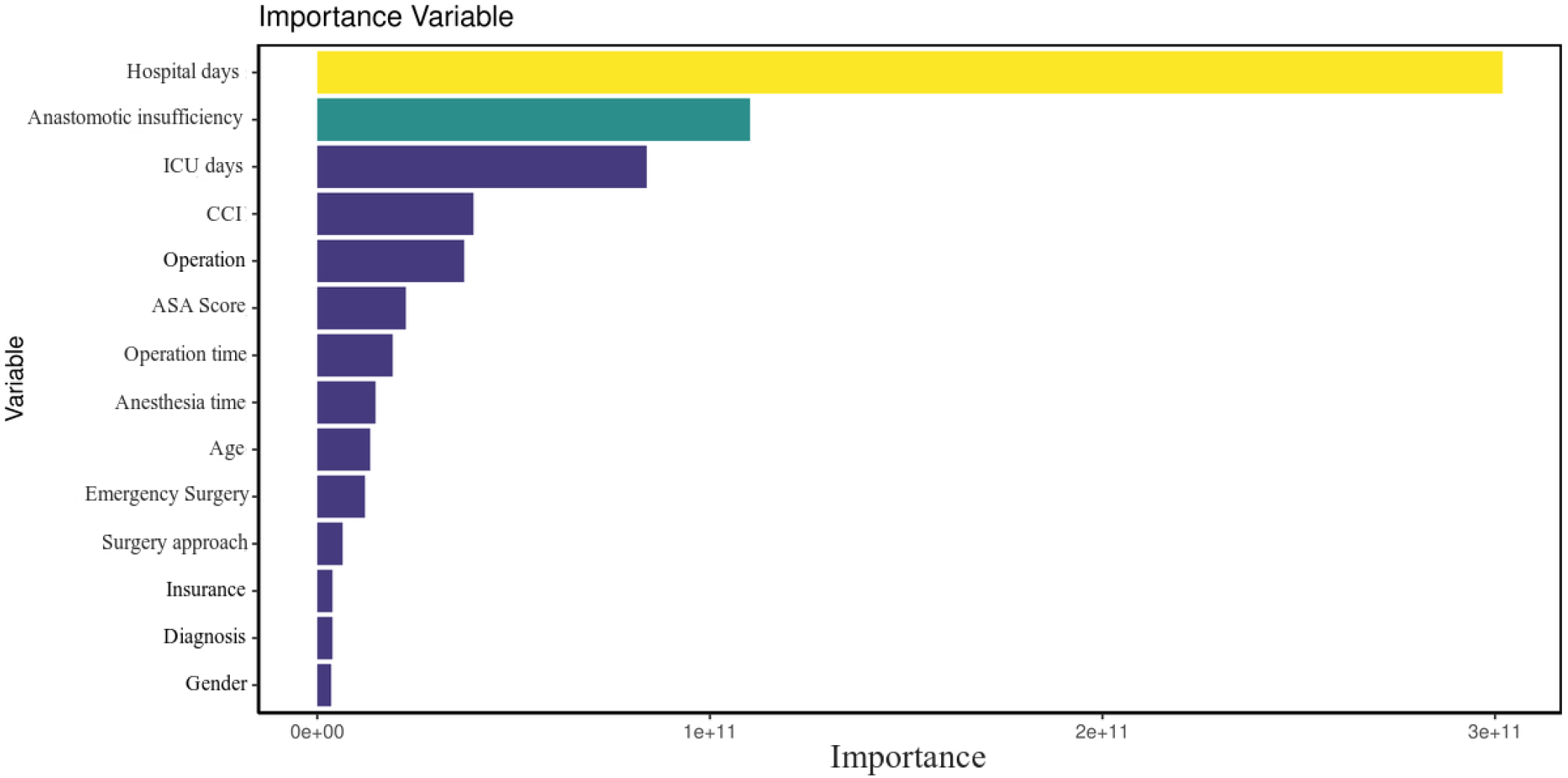
Total decrease in node impurities, measured by the Gini index from splitting on the variable, averaged over all trees.

**Figure 2.**
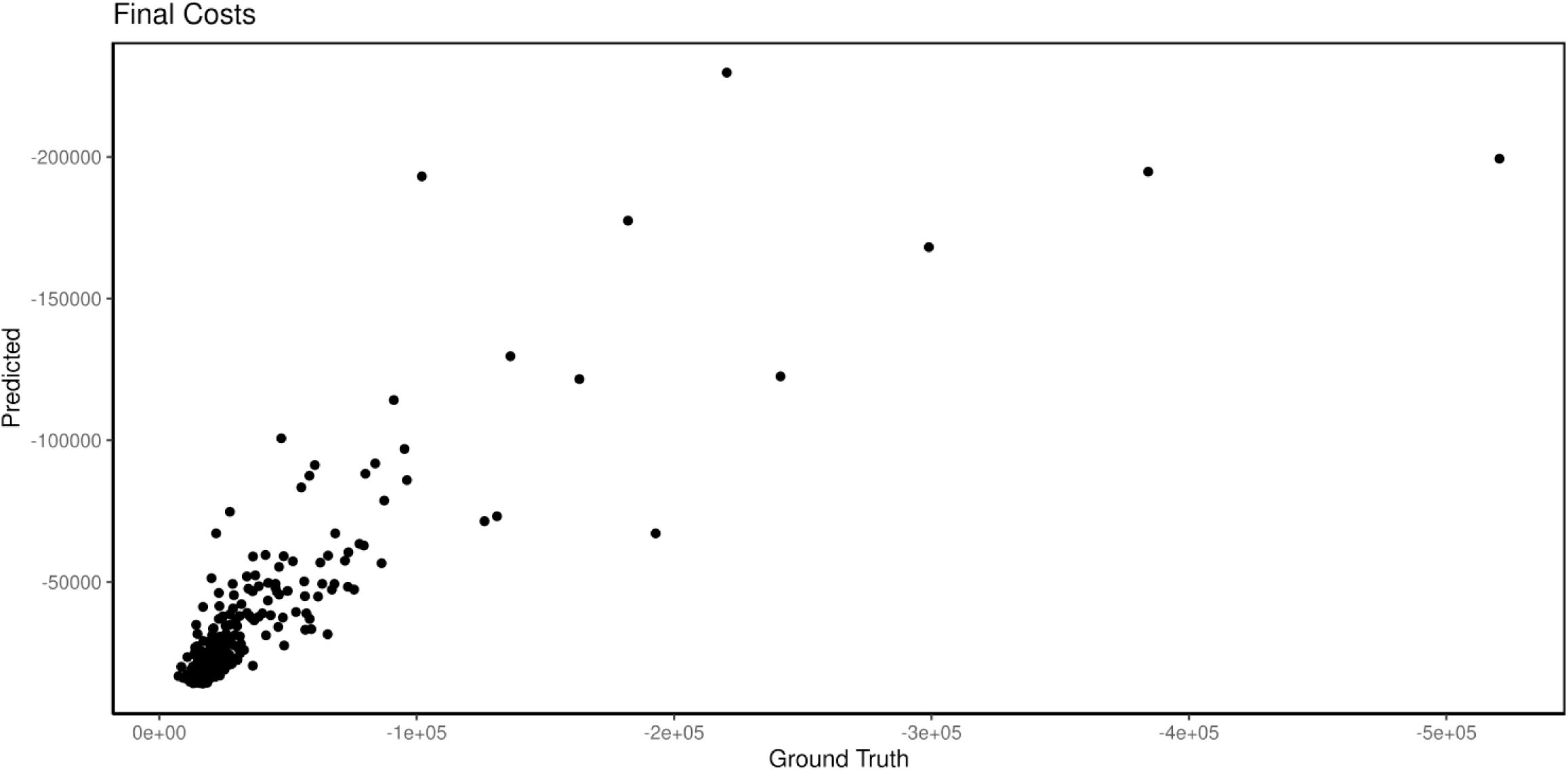
Predicted vs. Real observations of the model (Ground Truth).

In Figure 1, one can see those factors such as LOS and anastomotic insufficiency as well as intensive care unit stay are the best predictors in our model, which could be explained as being variables that are often correlated with post-operative complications and thus being more costly. The factor of hospitalization can be explained as being a good predictor of cost because the overall costs for a hospital will increase if the patient is not progressing after surgery. The same can be said about the intensive care unit. For the anastomotic insufficiency cases, it is evident as well that these complications bare a higher burden on the final costs. Mean decrease in Gini is the mean of a variable’s total decrease in node impurity, weighted by the proportion of samples reaching that node in each individual decision tree in the random forest. A higher mean decrease in Gini indicates higher variable importance. In other words, a node impurity is a measure of how much the model error increases when a particular variable is randomly permuted or shuffled.

Figure 2 indicates that the predicted values are not far off the actual observed values based on our data set. For most of the observations, our model was able to perform decently in predicting the final costs.

Figure 3 displays the Bland-Altman plot. The following information can be derived visually from the diagram: (1) an estimate of the true value on the x-axis (mean) (2) standard deviation (3) whether and to what extent systematic measurement errors (bias) lead to the deviations (variability was eliminated by difference formation on y-axis) (4) whether the deviation of the methods or the dispersion of the deviation depends on the level of the measured values (5) and whether outliers are present. Based on the plot one can imply that the values are mostly well distributed and not many outliers occur.

**Figure 3.**
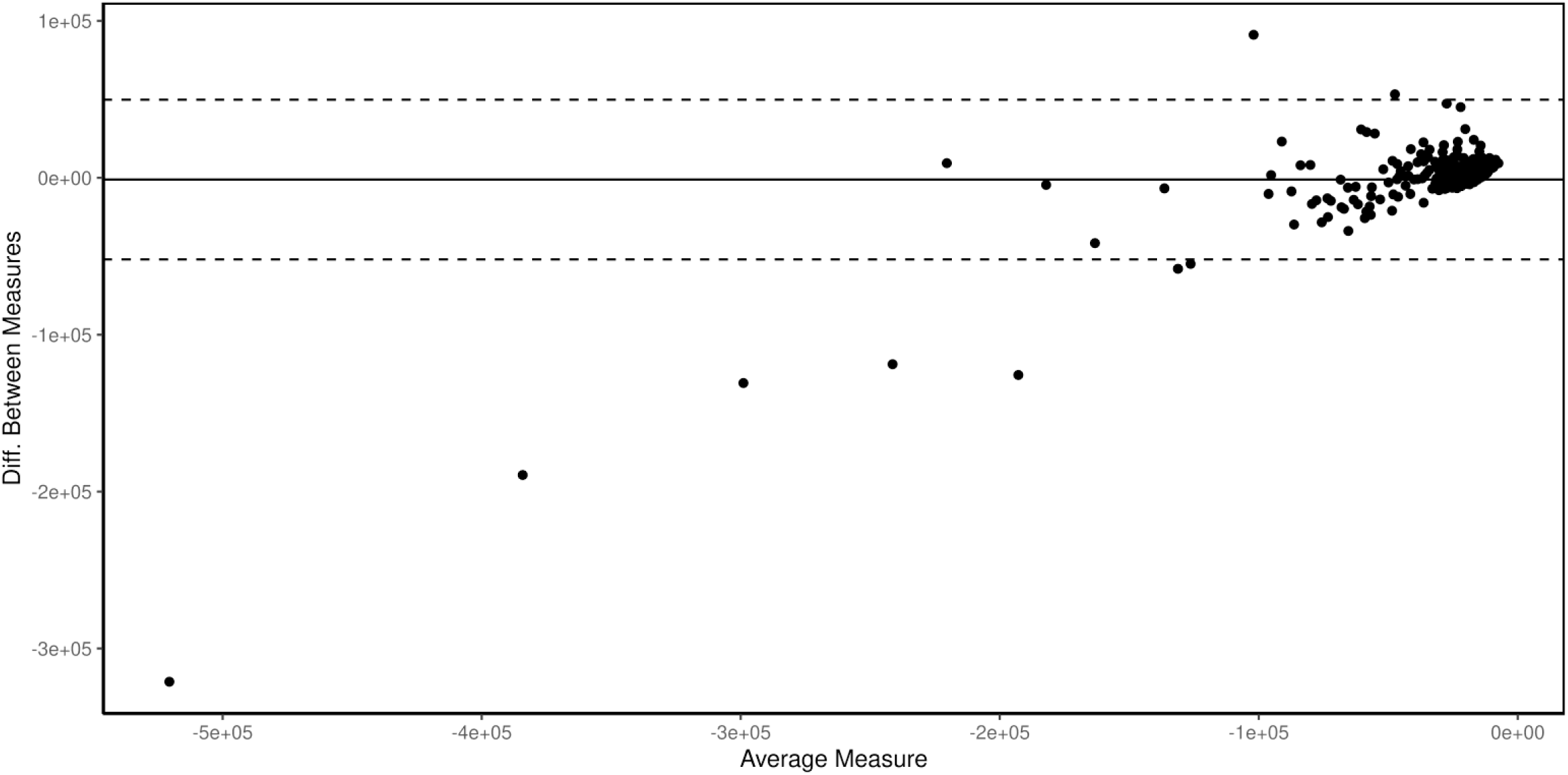
Bland-Altman Plot, measured by the difference of both measured values (S1 - S2) plotted on the Y-axis against the mean value (S1 + S2 /2) on the X-axis.

## 4. Discussion

Cost and finances play an increasingly important role in today’s healthcare system. It is imperative that hospitals control their costs more accurately beforehand and estimate the expenditure so that they do not get into financial difficulties.

Especially in surgery, and specifically colon surgery, this predictive model allows us to better manage and optimize the process in front of the surgeon and hospital.

### 4.1 Interpretation of results

As indicated, in this study, three models were developed and tested. The results show that random forest has the lowest percentage for all the costs examined on MAPE.

The lowest MAPE percentage for the random forest model indicates that this model is the most accurate at predicting costs associated with surgeries compared to the other two models examined. Typically, MAPE is a measure of error. It is used to measure the accuracy of a forecast (12). In calculating MAPE, the difference between the actual value and the forecast value is determined and expressed as a percentage. This means that if the difference between the actual value and the forecast value is small, the percentage is small (13). On the other hand, if the difference between the actual value and the forecast value is large, MAPE percentage is large. This implies that a small MAPE percent is an indication of the forecast value being near the actual value. In other words, the forecast value is more accurate (14). In the case of the three models, since the random forest model had the lowest MAPE percent value for all the costs compared to the other models considered, it is the most effective model in predicting the cost.

Why is Random Forest the most effective predictive model compared to the decision tree and generalized boosted regression models? This question can be answered by examining the model. The random forest model is a machine learning technique that is used to solve classification and regression problems (15). This model uses ensemble learning, a technique that combines many classifiers to obtain solutions to complex problems. A random forest algorithm comprises of multiple decision trees. The forest that is generated by the algorithm is trained through bootstrap aggregating or bagging (16). Bagging is a meta-algorithm that improves the machine learning algorithms’ accuracy.

The random forest algorithm establishes the result from the predictions of decision trees. It predicts by taking the mean of the prediction output of the various trees (17). This implies that the predicted outcome by the algorithm becomes more accurate when the number of decision trees is increased.

One of the features of the random forest model that makes it more accurate in predicting cost outcomes, is it reduces the overfitting problem normally experienced when using the decision tree model. As indicated, the model uses an ensemble learning method based on bagging (15, 18). In other words, the model creates many decision trees and then considers the outcomes of all the trees in its final prediction, enhancing the accuracy of the prediction by the model.

However, despite the higher accuracy of the random forest model when compared to the decision tree and generalized boosted regression models, the model does not have the highest possible accuracy when considered alone. Normally, when examining the accuracy of a prediction using MAPE, the result of less than 10% is considered highly accurate. A MAPE score of less than 20% denotes a good forecast, while that between 20 and 50% is considered a reasonable forecast (12). Looking at the results, it shows that the random forest model gives mostly reasonable forecasts rather than accurate forecasts. The model gave an outcome of over 20% when analyzed using the MAPE. This means that while it is the most accurate model when compared to the other models, when considered alone, it has only considerable accuracy and it does not accurately predict the cost incurred.

A number of similar studies have been carried out on the random forest model in terms of its accuracy in predicting outcomes. For example, Mei et al. (2014) examined the prediction accuracy of the random forest model when applying real-time forecasting of the New York electricity market (18). In reviewing the model’s prediction accuracy, its results were compared to that of the auto-regressive-moving-average model and an artificial neural network model. It was established that the random forest model exhibited a lower MAPE value. The results in the study by Mei et al. (2014) are similar to those of this study which also show that the random forest model has a higher level of making fewer mistakes by predicting when compared to other studies (18). However, the shortcoming of the study by Mei et al. (2014) is it compares the random forest model to only two other models. This does not provide adequate insight into the model’s prediction accuracy (18). A comparison with additional models would have helped determine whether the random forest model was the most accurate prediction model or if others were more accurate.

Another similar approach to comparing algorithms was made by Xu et al. (2021), which developed and tested an accurate prediction model based on the random forest classification algorithm (19). They evaluated the prediction for inland water quality. To evaluate the performance of the model, the researchers compared it to other models: multilayer perceptron, SVR (support vector regression), KNN (K-Nearest Neighbor), ridge regression, gradient boosting regression, bagging and decision tree. It was established that the random forest-based prediction model had the highest level of accuracy when compared to all the other prediction models examined. This implies that random forest provides the most accurate outcomes when used for prediction. The results in the study by Wang et al. (2020) align with those of this study since it was also established that the random forest model is the most accurate compared to other models. The study by Wang et al. (2020) provides better insight into the accuracy of the random forest model because it compared it to multiple models (19). It is an indication that the random forest model is one of the most accurate prediction models that can be used to predict costs for surgery.

Lastly, the results are in line with those of Toqué et al. (2020), who also established that the random forest model has a higher accuracy compared to other models (20). In the study, Toqué et al. (2020) built and tested machine learning models for forecasting the Montreal subway smart card entry logs using event data to find an optimal model that accurately predicts the number of incoming passengers at each station of a transportation network (20). The prediction models were random forest, gradient boosting decision trees, artificial neural networks and kernel-based models, including a support vector regressor and a gaussian process (20). The results showed that all the random forest models performed best using RMSE for the evaluation, and did decent using MAPE and MAE.

The results in this study show that all the models have reasonable accuracy as the MAPE for each of all the costs highlighted is below 50%. This means that all of the models can be used to predict the costs to some level of accuracy. However, when compared it can be seen that the random forest model is a more accurate predictor. These results are evident in similar studies showing that the random forest model is a more accurate prediction model.

### 4.2 Implications

One of the implications of the results is that hospitals and other concerned parties can employ the random forest model to forecast costs not only for colon surgery but also the costs of other risks and conditions mentioned previously. This work lays the foundation for further work and research in this area. This will allow for better financial calculations for hospitals. Through such a predictive model, it is possible to better estimate medical costs, which is especially important when factors such as LOS in the hospital and ICU as well as complications such as anastomotic insufficiency can have a large financial impact on the high cost. The results show that the random forest model provides more accurate predictions compared to other models like generalized boosted regression and decision tree models. It means that for concerned parties to achieve more accurate results when predicting the costs of conditions or any other outcome, the random forest model should be employed.

Another implication is that there is a need for further research about the model in terms of enhancing the accuracy of the random forest model. The results show that for the final costs examined, the accuracy is more than 20%. This is only reasonable accuracy. However, it is way before the desired value. As indicated, the MAPE value of less than 20% is an indication of a good forecast, while that of less than 10% shows that the forecast is highly accurate. While achieving a highly accurate forecast is unlikely, any good prediction model should give a good forecast. With the random forest model being the most accurate model, this implies that it should be developed further to improve accuracy as to give more credible results when used to predict outcomes, meaning further research is needed on the model.

Despite the good implications and the wide range of applications, the ethical aspect should not be ignored. Naik et al. have shown in their work that there are currently no well-defined guidelines when it comes to treating people with an application such as this. They mention that transparency must be created when working with such algorithms. Furthermore, weaknesses such as cyberattacks and privacy invasion should not be ignored if you want to advance this field and research (21).

### 4.3 Limitations of the study

The main limitation of the study is a lack of a representative sample. In this case, the focus was on patients undergoing colon surgery. However, in the sample dataset, only 347 individuals met this criterion. This implies that the sample was not selected in the manner that made it representative of patients undergoing colon surgery. The larger the dataset, the more accurate the results are. However, the limited number of individuals with common reasons for higher costs implies that it was not possible to effectively test the developed models in terms of their ability to predict costs associated with the disease. For such models, there is a need for adequate and detailed data to ensure they are tested thoroughly. Additionally, an overall increase in the sample size could result in more precise models by looking at the values in Table 4. Especially the events per predictor should be bigger.

**Table 4.** Internal validation performance for the 3 developed models

| Classifier | MAPE (%) Final Costs |
| --- | --- |
| Random Forest | 21.4 (17.2-26.8) |
| Decision Tree | 25.2 (21.4-26.3) |
| Generalized Boosted Regression | 29.7 (25.2-34.2) |

Note: Scores Reported as Mean (95% Confidence Interval)

## 5. Conclusion

Post-operative complications such as anastomotic insufficiency, ICU or hospital LOS increase the cost burden for patients and hospitals. Also, preoperative conditions like CCI increase cost. However, there is no way of predicting these costs so that a patient or healthcare system can prepare adequately to handle the condition. This study thereby aimed to develop and validate a prediction model to accurately predict cost and develop strategies to eliminate or cover them. Out of the three tested models, the results obtained based on MAPE analysis showed that the random forest model is the most accurate. Therefore, the results imply this model should be adopted for prediction. However, the fact that MAPE results were mostly over 20% means that further research should be undertake on improving its accuracy.

## Data Availability

All data produced in the present study are available upon reasonable request to the authors

## Author Contributions

Conceptualization A.T., B.E., data collection B.E.; analysis V.O., A.T.; visualization A.T., writing—original draft preparation A.T, V.O.; writing—review and editing D.F., M.D.H., P.C.C. All authors have read and agreed to the published version of the manuscript.

## Data Availability Statement

The datasets used and/or analysed during the current study are available from the corresponding author on reasonable request.

## References

1. Xi, Y. and Xu, P., 2021. Global colorectal cancer burden in 2020 and projections to 2040. Translational Oncology, 14(10), p. 101174.

2. Soeters, P.B., de Zoete, J.P.J.G.M., Dejong, C.H.C., Williams, N.S. and Baeten, C.G.M.I., 2002. Colorectal surgery and anastomotic leakage. Digestive surgery, 19(2), p. 150.

3. Karliczek, A., Harlaar, N.J., Zeebregts, C.J., Wiggers, T., Baas, P.C. and Van Dam, G.M., 2009. Surgeons lack predictive accuracy for anastomotic leakage in gastrointestinal surgery. International journal of colorectal disease, 24(5), pp. 569–576.

4. Kourou, K., Exarchos, T.P., Exarchos, K.P., Karamouzis, M.V. and Fotiadis, D.I., 2015. Machine learning applications in cancer prognosis and prediction. Computational and structural biotechnology journal, 13, pp. 8–17.

5. Mosavi, A., Ozturk, P. and Chau, K.W., 2018. Flood prediction using machine learning models: Literature review. Water, 10(11), p. 1536.

6. Musunuri, B., Shetty, S., Shetty, D., Vanahalli, M., Pradhan, A., Naik, N., Paul, R., 2021. Acute-on-Chronic Liver Failure Mortality Prediction using an Artificial Neural Network. Engineered Science, 15, pp. 187–196.

7. >Hameed, B.M.Z.; S. Dhavileswarapu, A.V.L.; Raza, S.Z.; Karimi, H.; Khanuja, H.S.; Shetty, D.K.; Ibrahim, S.; Shah, M.J.; Naik, N.; Paul, R.; Rai, B.P.; Somani, B.K. Artificial Intelligence and Its Impact on Urological Diseases and Management: A Comprehensive Review of the Literature. J. Clin. Med. 2021, 10, 1864.

8. Collins, G.S., et al., Transparent reporting of a multivariable prediction model for individual prognosis or diagnosis (TRIPOD): the TRIPOD statement. 2015. 102(3): p. 148–158.

9. Bolenz, Gupta, Roehrborn, Lotan, Predictors of costs for robotic assisted laparoscopic radical prostatectomy, Urologic Oncology: Seminars and Original Investigations, Volume 29, Issue 3, 2011, Pages 325–329.

10. Abidin, S. and Jaffar, M.M. (2014) Forecasting Share Prices of Small Size Companies in Bursa Malaysia. Applied Mathematics and Information Sciences, 8, 107–112

11. Sushmita, Newman, Marquardt, Ram, Prasad, De Cock, Teredesai, 2015. Population Cost Prediction on Public Healthcare Datasets. In Proceedings of the 5^th^ International Conference on Digital Health 2015. Association for Computing Machinery, New York, NY, USA, 87-94.

12. Coleman, C.D. and Swanson, D.A., 2007. On MAPE-R as a measure of cross-sectional estimation and forecast accuracy. Journal of Economic and Social Measurement, 32(4), pp. 219–233.

13. Hyndman, R.J. and Koehler, A.B., 2006. Another look at measures of forecast accuracy. International journal of forecasting, 22(4), pp. 679–688.

14. Rayer, S., 2007. Population forecast accuracy: does the choice of summary measure of error matter?. Population Research and Policy Review, 26(2), pp. 163–184

15. Sinha, P., Gaughan, A.E., Stevens, F.R., Nieves, J.J., Sorichetta, A. and Tatem, A.J., 2019. Assessing the spatial sensitivity of a random forest model: Application in gridded population modeling. Computers, Environment and Urban Systems, 75, pp. 132–145

16. Bharathidason, S. and Venkataeswaran, C.J., 2014. Improving classification accuracy based on random forest model with uncorrelated high performing trees. Int. J. Comput. Appl, 101(13), pp. 26–30.

17. Wang, L., Liu, Z.P., Zhang, X.S. and Chen, L., 2012. Prediction of hot spots in protein interfaces using a random forest model with hybrid features. Protein Engineering, Design & Selection, 25(3), pp. 119–126.

18. J. Mei, D. He, R. Harley, T. Habetler and G. Qu, “A random forest method for real-time price forecasting in New York electricity market,” 2014 IEEE PES General Meeting | Conference & Exposition, 2014, pp. 1–5.

19. Xu, J.; Xu, Z.; Kuang, J.; Lin, C.; Xiao, L.; Huang, X.; Zhang, Y. An Alternative to Laboratory Testing: Random Forest-Based Water Quality Prediction Framework for Inland and Nearshore Water Bodies. Water 2021, 13, 3262.

20. Toqué, F., Côme, E., Trépanier, M., Oukellou, F., 2020. Forecasting of the Montreal subway smart card entry logs with event data. TRB, CIRRELT-2020-33.

21. Naik N, Hameed BMZ, Shetty DK, Swain D, Shah M, Paul R, Aggarwal K, Ibrahim S, Patil V, Smriti K, Shetty S, Rai BP, Chlosta P and Somani BK (2022) Legal and Ethical Consideration in Artificial Intelligence in Healthcare: Who Takes Responsibility? Front. Surg. 9:862322.

